# Tracking the Covid-19 pandemic : Simple visualization of the epidemic states and trajectories of select European countries & assessing the effects of delays in official response

**DOI:** 10.1101/2020.03.14.20035964

**Authors:** A. Kévorkian, T. Grenet, H. Gallée

## Abstract

We present a self-synchronizing and robust method for comparing the progression of the Covid-19 epidemics among multiple countries. In their growth phase the epidemics show power law rather than exponential law time dependences. They are similar enough for the earlier China outbreak to guide other countries projections. The delayed reaction of European countries is shown to produce a significantly worse outcome compared to China.

## Introduction

As the Covid-19 disease is currently spreading across many countries around the world, it is of crucial importance for public authorities to be able to easily estimate the dynamics of the epidemic, the expected effects of restrictions imposed on the population, and the impact of delays in their application.

China suffered from the epidemic outbreak earlier than the rest of the world and imposed immediate and stringent confinement measures as a reaction. The positive effects of the Chinese measures have become evident in the past few weeks, and the Chinese data can thus be used as a reference to better analyze the situation in other countries. A variety of approaches may be used. One example consists in collecting data on various cities in a given country and comparing them to a set of historical data collected in a series of cities judged relevant in China. Coupled with epidemic spreading modeling, this procedure may help extrapolate the trends to be expected in the cities under study, and to deduce forecasts for the entire country. Unfortunately, as the European cities under consideration are still in a very early stage of the epidemic’s development, this approach is very imprecise and the extrapolations obtained may be of weak reliability. For instance, one such study released on March 8, 2020 predicted that the final total number of diagnosed confirmed cases in Italy after the end of the epidemic would reach approximately 7,000 +/- 1,000 [1]. Today we know that this number was significantly underestimated, as four days later the official records exceeded 15,000 confirmed cases. This shows the need of a simpler but more robust method in order to guide authorities.

In this short note, we suggest a simple way of visualizing the epidemic trajectories of select countries to enable an easy and direct comparison with China. It immediately appears that several European countries have followed rather closely China’s trajectory up until now.

Order of magnitude estimates of what may be expected for the weeks to come can thus be provided. The estimates provided below highlight the importance of rapid authority response time. Simply stated: in the race against the clock, any delays in authority reaction will allow for the further spread of the epidemic.

## Method and analysis

In figure 1, rather than time series, we plot for each day the number of daily new confirmed cases as a function of *N*, the total number of confirmed cases since the onset of the epidemic, for several countries [2,3]. This is equivalent to phase space plots commonly used to represent dynamical systems in Physics. While each dot represents the data from a given day, time does not explicitly appear. Such a plot enables a direct comparison of the otherwise nonsynchronous curves of the disease progression for the different countries.

**Figure 1:**
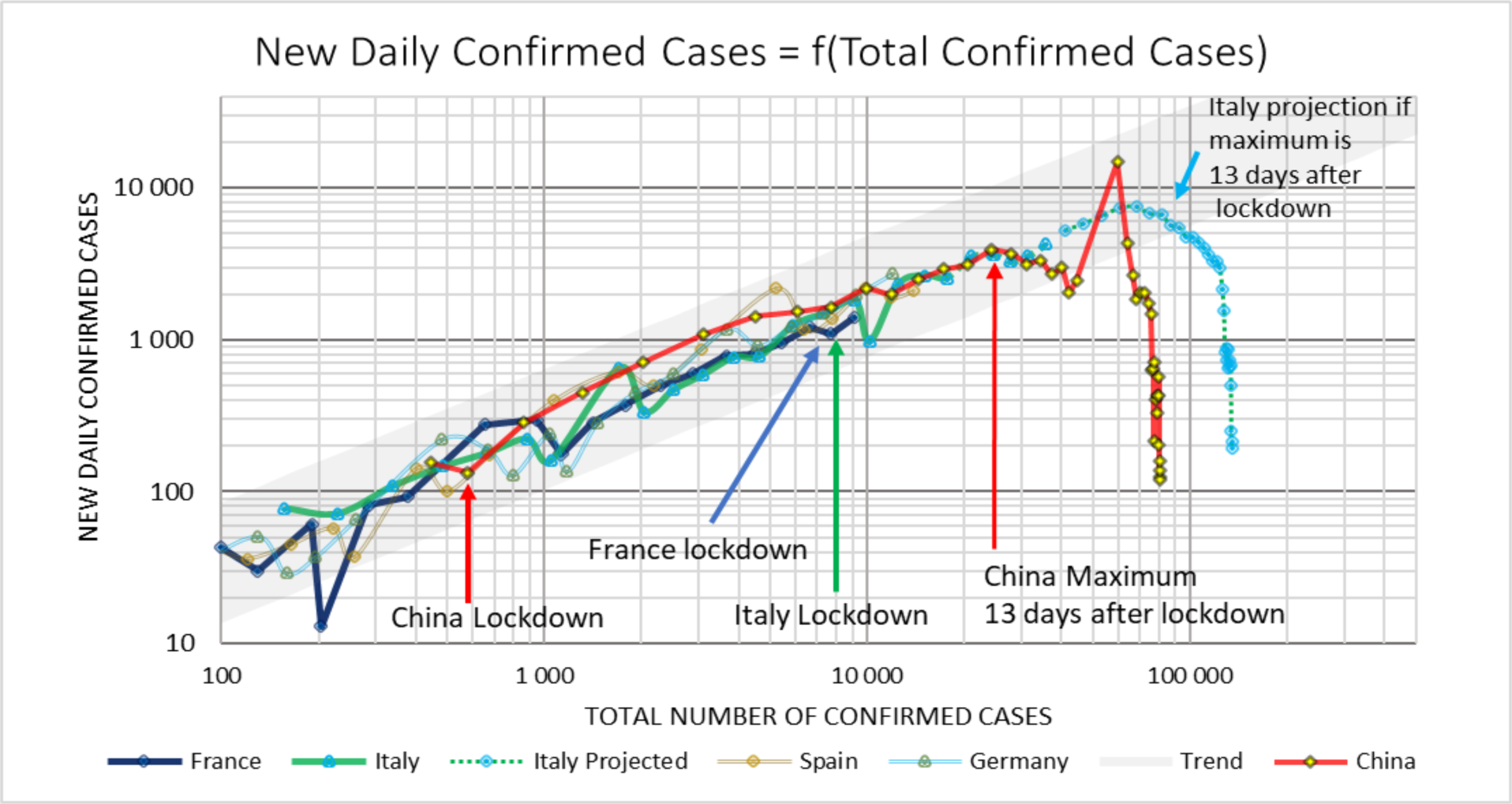
Number of new daily confirmed cases as a function of *N*, which is the total number of confirmed cases since onset of Covid-19 epidemic for China, Italy, France, Germany and Spain. Each point corresponds to a specific day. All of the countries exhibit similar epidemic progression within a certain band. Projection for Italy (dotted line and blue points) takes into account a 13-day delay beyond lockdown to reach the maximum daily confirmed cases as observed in China. The sharp peak on China’s curve is due to a change of counting method on Fed 12 and is discussed in Appendix 1.

**Fig. 2 :**
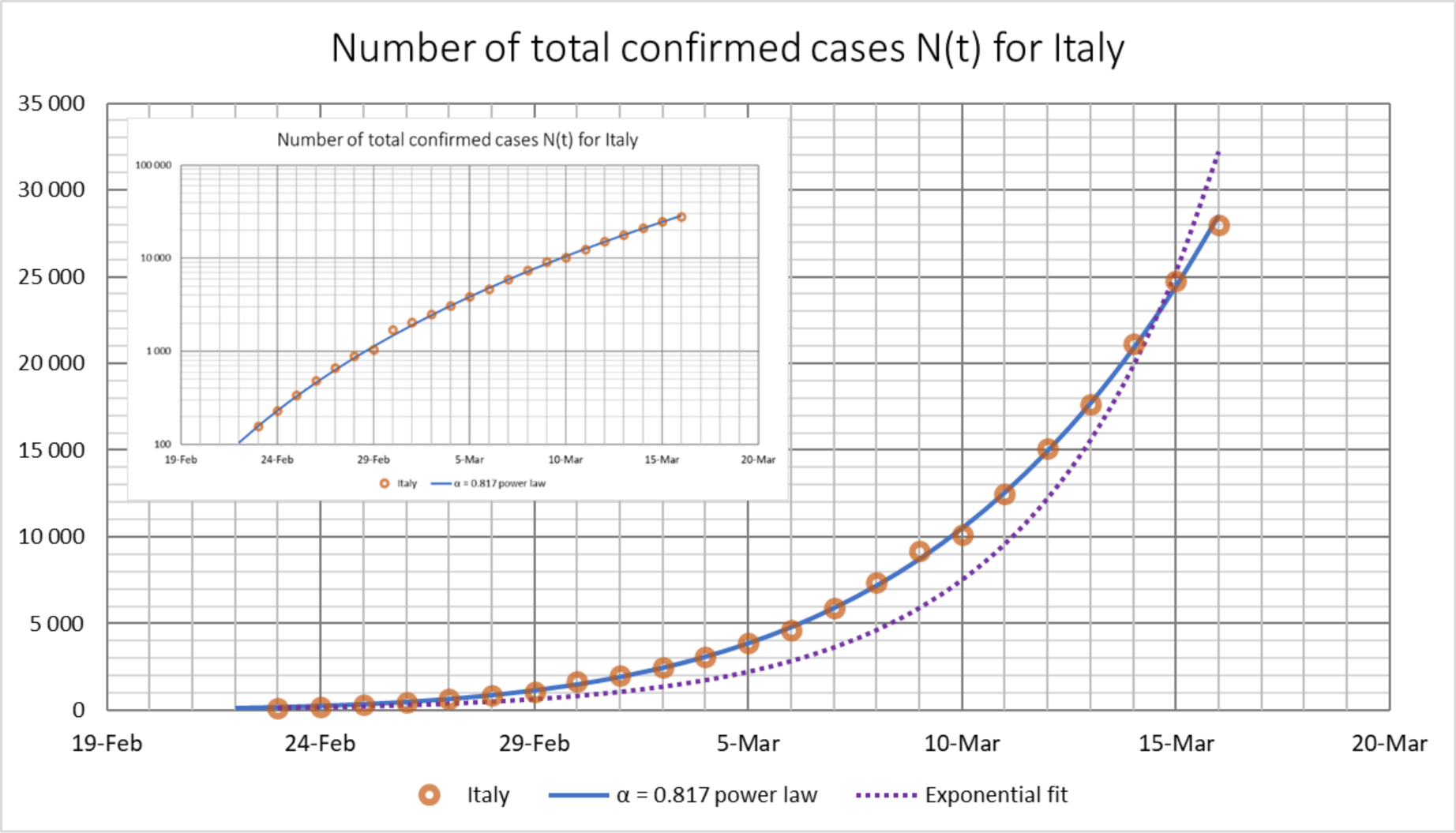
The main graph represents the total number of confirmed cases data points vs time (red circles), in close agreement with the power law from eq. (1) of main text (blue curve) and differs markedly from the best exponential fit (dotted curve, least square error fit). Insert: semi-log plot of *N*(*t*) showing the adequacy of eq. (1) at all orders of magnitude.

A further feature of this plot is that it is largely independent of the case detection methods which are known to vary both from country to country as well as over time for a given country. When a specific country decides to change its detection strategy, a perturbation appears transiently on the curve and then is rapidly dampened in the following days. In other words, the plot outlines possible measurement inconsistencies and at the same time provides the underlying trend of the epidemic for a given country. A detailed analysis of these benefits is given in Appendix 1.

As data span several orders of magnitude, logarithmic scales are used. It is clearly visible that within individual variations and data collection rule modifications, the China trajectory is tracked rather closely by the other countries.

A key observation is that the maximum of the daily confirmed cases in China occurred some 13 days after a stringent lockdown was put into place in Wuhan and its region. This 13-day delay is similar to the one separating the onset of symptoms from the time of diagnosis, as found in interviews of patients [4]. Beyond the maximum, it is worthwhile noting that the China curve falls off as the number of daily new cases starts to decrease. The number of confirmed cases continues to increase; nevertheless at a slower pace, and finally stabilizes at a significantly higher value than what was recorded at the maximum. At this stage, the number of daily cases has become very small and the virus propagation is essentially halted.

A comparison of plots in Figure 1 shows that the Italian lockdown versus the Chinese lockdown was ordered approximately 10 days later in the epidemic progression. One may readily try to estimate the final number of cases to be expected by assuming that the Italian lockdown will have the same effects as the Chinese lockdown. The ascending part of the Italian curve can be approximated by a straight line with a slope *α* = 0.817 in logarithmic scale. As shown in Appendix 1 this corresponds to the following time dependence for the total number of confirmed cases since epidemic onset *N*(*t*):

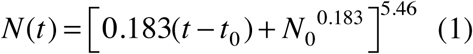

where *N(t)* is the number of confirmed cases as a function of time expressed in days starting on February 23, 2020 with *N*_0_ = 157 confirmed cases. The dynamics is thus described by a power law, rather than by an exponential law. Assuming this progression regime remains the same across the entire ascending part of the curve, the maximum number of confirmed cases for Italy is estimated to reach around 55,000. Assuming further the efficiency of the Italian lockdown to be similar to that of China, we then add a downward slope matching the Chinese one. This triggers an additional number of cases equal to 1.7 times the number of cases at the maximum, yielding a final number reaching approximately 140,000 confirmed cases for Italy.

A precise estimation of the final death toll that can be expected is outside the scope of this note. Calculating this would require knowledge of the actual mortality rate; e.g. the final number of deaths relative to the final number of infections. If symptomatic cases are the only ones counted, then the number of infection cases are underestimated, and the apparent mortality rate is computed to be higher than it actually is. During the epidemic progression this apparent mortality rate may change over time. Since the epidemic onset in Italy for instance, as of March 15 a steady increase in the apparent mortality rate can be observed. This may be due to a combination of causes: a decrease in the efficiency of case detection as the number of infections grows, a concomitant decrease in medical care efficiency, and effects linked to the time-lag between the detection of early symptoms and actual death [5]. If one uses the WHO’s average mortality rate estimate of 3.4% then Italy’s final death toll would rise to approximately 5,000. However with the significantly higher apparent mortality rate of 8.3% observed on March 18, 2020 [2] and its still increasing trend, more than 10.000 deaths may be expected. This range of numbers should be compared to the approximately 3,300 deaths in China.

In the case of France, school and University closure decisions were announced on March 12, 2020 by president Emmanuel Macron, with strict confinement measures effective March 17, 2020, which is close to the same point in epidemic progression as when Italy applied full lockdown on March 9, 2020. The French case may lead towards an outcome in number of cases of the same magnitude as Italy’s, with a smaller final death toll as the observed apparent mortality is smaller as of March 18. It currently appears that the France is moving in a direction comparable to Italy’s in terms of the death toll, with France nevertheless appearing to have a lower mortality rate as of March 18, 2020.

Our approach thus provides guideline estimates based on the currently available data. It demonstrates how sharply the confirmed cases and the corresponding death toll will increase, as both France and Italy waited longer than China before taking lockdown actions. For Italy alone, the 10-day lag compared to China may engender approximately three times more deaths. In addition, Germany and Spain appear to be at a similar stage in epidemic progression, indicating that the total death toll in Europe will far exceed China’s death toll.

It should be stated that the data coming from China was made available early to European authorities, and thus allowed for more time to prepare an informed and coordinated response to the coming outbreak. The earlier lockdown in China appears to have significantly limited the impact of the Covid-19 epidemic there. The time lost in European countries to initiate a similar level of action may unfortunately result in a death toll increased by the thousands relative to the China outbreak.

## Conclusion

We have introduced a “phase space plot” of the covid-19 epidemic progression where, instead of time series, the number of daily new cases is plotted as a function of the total number of confirmed cases since epidemic onset. This representation makes clear that the countries considered, within fluctuations, follow very similar epidemic trajectories. It allows an easy extraction of the mathematical law governing the expansion phase and, using the China data as a guiding example, suggests a way to estimate the final outcome of lockdown timing and its strength.

The evolution of the characteristic time for doubling the daily confirmed cases is often used to measure the speed of epidemic progression. If the epidemic dynamics law was exponential one would expect the doubling time to be constant in time. In the countries considered here the epidemic progression is not exponential but of power law type, which implies an in-built gradual increase of the doubling time. Therefore this increase does not prove by itself that sanitary measures are being effective. Such a conclusion can only be reached when a change of dynamical regime occurs, showing a persistent deviation from the trend such as shown in Fig.1 or from the law given by eq. (1) for Italy and eq. (3) of appendix 1 for other countries (with appropriately determined values of *α*).

In this short note we have only exemplified a few European countries and mainly focused on Italy as an example, but our approach may be applied to other countries as well. We hope that visual representations like the one we present in this note can help better anticipate the Covid-19 epidemic dynamics and provide a crucial tool for public officials’ decision making.

## Data Availability

All data used are public and available through the internet links provided in the paper.

## Appendix 1

Straight lines with slope *α* in the log-log plot of figure 1 are solutions of the differential equation:

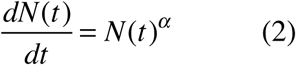

where the time derivative of *N*(*t*), the total number of cases since epidemic onset, represents the daily variation of cases.

Unless when *α* = 1 which corresponds to an exponential *N*(*t*), this equation has solutions of the type:

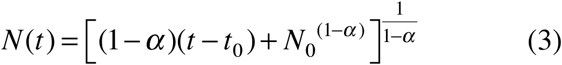

With *α* = 0.817 one gets eq. (1) of the main text. Any couple (*t*_*0*_, *N*_*0*_) may be used as long as *N*_*0*_ is large enough to be devoid of statistical fluctuation. As a direct check we show in Fig. 2 below that the observed *N*(*t*) for Italy indeed follows this law very closely.

Most measuring methods will underestimate the actual number of cases *N*_*act*_ (*t*), which we can describe using an unknown proportional constant *K* such that *N*_*act*_ (*t*) = *KN*(*t*).

Suppose now that *N*_*act*_ (*t*) follows equation (2). Then 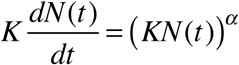. Taking the logarithm, one obtains: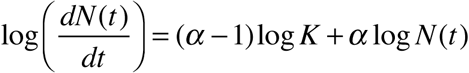. Since *α* is close enough to 1 and *K* of the order of a few units: |*α* − 1| log *K* ≪ *α* log *N* (*t*) and 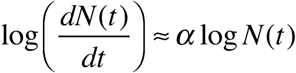. Using the measured *N*(*t*) to construct the phase space plot of Fig. 1 thus gives the same slopes as if the actual *N*_*act*_(*t*) was used.

Now assume that a country suddenly changes its counting method, resulting in *K* changing to *K*_*new*_: the corresponding points on the country curve will be shifted vertically. But as new cases counted with *K*_*new*_ accumulate, they will outnumber the previous points and the curve will revert to the underlying trend (within a gradually smaller vertical shift) and to its associated slope. An example is seen in China’s curve in Fig. 1. The sharp peak corresponds to the country’s decision of February 12 to extend counting to “clinically diagnosed cases” (e.g. using CT scans), a decision that suddenly increased the numbers and provoked some confusion. It only alters our plot locally.

Finally it can be checked that if the efficiency of case counting gradually decreases with time for a country, which may be the case as suggested by the increase of apparent mortality observed in several countries (see main text), curves in Fig. 1 are only slightly altered.

## References

[1] Prediction of COVID-19 Spreading Profiles in South Korea, Italy and Iran by Data-Driven Coding, Choujun Zhana, Chi K. Tseb, Zhikang Laic, Tianyong Haoa and Jingjing Su, https://www.medrxiv.org/content/10.1101/2020.03.08.20032847v1.full.pdf

[2] Data were taken from https://www.worldometers.info/coronavirus/ and references therein

[3] All numbers shown are officially confirmed values. It is known that the actual number of cases may be larger by factors depending on the way the various countries proceed to detect cases.

[4] Characteristics of and Important Lessons From the Coronavirus Disease 2019 (COVID- 19) Outbreak in China: Summary of a Report of 72 314 Cases From the Chinese Center for Disease Control and Prevention, Wu Z and McGoogan JM., JAMA. Published online February 24, 2020. doi:10.1001/jama.2020.2648

[5] Real estimates of mortality following COVID-19 infection, D. baud, X. Qi; K. Nielsen-Saines, D. Musso, Léo Pomar and G. Favre, Lancet (2020) published online March 12, https://doi.org/10.1016/S1473-3099(20)30195-X

